# Association of APOE gene polymorphisms and multiple risk factors with hypertension in middle-aged and elderly Tibetan population in Tibet on Bayesian network model

**DOI:** 10.1101/2024.11.18.24317520

**Authors:** Yufei Zhang, Jing Zhang, Yuan Yao, Jiaojiao Yan, Hua Chun, Hai Xiong

**Author notes:** **Corresponding Author: Hua Chun:**, Medical College of Tibet University, No. 10, Zangda East Road, Chengguan District, Lhasa, 850000, Tibet Autonomous Region, China; **Hai Xiong:**, Medical College of Tibet University, No. 10, Zangda East Road, Chengguan District, Lhasa, 850000, Tibet Autonomous Region, China; Hua Chun and Hai Xiong are Co-corresponding authors.

## Abstract

**BACKGROUND:** The distribution of Apolipoprotein E (APOE) gene and its relationship with hypertension at high altitude in Tibet have not been elucidated. The purpose of this study was to understand the distribution of APOE genotypes and APOE rs7412, rs429358 genotypes in the middle-aged and elderly Tibetan population in Tibet, and to explore the related influencing factors by constructing a Bayesian network model of hypertension in middle-aged and elderly Tibetans.

**METHODS:** From November 2021 to November 2022, a multi-stage cluster random sampling method was employed to collect data from 511 middle-aged and elderly Tibetans residing in the Ali region, Nagqu City, and Shannan City of Tibet. The APOE gene, rs429358 and rs7412 single nucleotide polymorphisms (SNPs) were analyzed by fluorescence polymerase chain reaction (PCR). Multivariate logistic regression analysis and Bayesian Network (BN) were applied to examine the relationship between Covariates and hypertension.

**RESULTS:** Among all subjects, the frequencies of genotypes ε2/ε3, ε2/ε4, ε3/ε3 and ε3/ε4 were 14.8%,1.8%,75.0%, and 8.4%, respectively. APOE gene rs7412 locus CC, CT and TT genotype frequencies were 83.4%, 0 and 16.6%, respectively, and rs429358 locus TT, TC and CC genotypes, genotype frequencies were 89.8%, 10.2% and 0. The results of this study indicated no correlation between APOE genotypes, APOE rs7412, rs429358 genotypes, and the prevalence of hypertension in middle-aged and elderly Tibetan population in Tibet. Sex, age, overweight obesity, hyperhomocysteinemia (HHcy) and High Altitude Pulmonary Edema (HAPC) represent direct risk factors for hypertension based on a Bayesian network.

**CONCLUSIONS:** APOE genotype and APOE rs7412, rs429358 genotypes were not genetically correlated with hypertension in middle-aged and elderly Tibetan populations in Tibet. The Bayesian network model revealed the direct and indirect factors and the strength of the association of the hypertension. Additionally, it elucidated the complex network relationships among these factors, which provided a scientific basis for the early prevention of hypertension among middle-aged and elderly Tibetan populations in Tibet.

## INTRODUCTION

More than 1 billion people worldwide suffer from high blood pressure^[1]^. High blood pressure is one of the world’s leading risk factors for death and disability. The number of people living with hypertension doubled between 1990 and 2019, from 650 million to 1.3 billion^[2]^. Currently, among adults aged 30–79 years with hypertension, only 54% have been diagnosed with the condition, 42% are being treated for their hypertension, and 21% are considered to have their hypertension controlled^[3]^. In 2015, the prevalence of hypertension among adult Chinese residents was 25.2%^[4]^, and 27.9% in 2018^[5]^. Several general environmental exposures can also contribute to increases in blood pressure, including pollution^[6]^, very cold temperatures and extreme elevations in altitude^[7]^. However, the main contributors seem to be personal environmental exposures such as poor-quality diet high in sodium and low in potassium, overweight and obesity, consumption of alcohol, use of tobacco and physical inactivity^[8]^.

The Tibet Autonomous Region is located in southwestern China, with an average altitude of over 4,500 meters above sea level, and the Tibetan population accounts for approximately 90% of the total. The prevalence of many chronic diseases, including hypertension, is higher than the national level, particularly among middle-aged and elderly populations^[9]^. The prevalence of hypertension in Tibet is approximately 37.6%^[10]^, while in other municipalities within the Tibetan region, it ranges from 32.5% to 62.4%^[11, 12]^, all of which exceed the national average prevalence.

The APOE gene is located on chromosome 19 and encodes three alleles, ε2, ε3 and ε4, which have been shown to affect the lipid profile and the development of coronary artery disease^[13, 14]^. The distribution of these alleles varies between ethnic groups: Europeans and African-Americans have a high frequency of ε4; Asians have a low frequency of ε2 and ε4^[13]^. There are two common single-nucleotide polymorphisms (SNPs) of the APOE gene: 526C > T(rs7412) and 388T > C (rs429358).

In this study, we detected the APOE genotype and the APOE rs7412 and rs429358 genotypes in the middle-aged and elderly Tibetan population. Additionally, we explored the influencing factors on the prevalence of hypertension among middle-aged and elderly Tibetans by constructing a Bayesian network model.

## METHODS

### Data Availability

The data that support the findings of this study are available from the corresponding author upon reasonable request.

### Study Design and Population

All participants completed a detailed questionnaire, including demographics, lifestyle and hypertension-related information. In addition, participants’ clinical information such as weight, height, and blood pressure were measured by trained staff. The following parameters, namely, Blood uric acid (UA), total cholesterol (TC), triglycerides (TG), low-density lipoprotein (LDL-C), high-density lipoprotein (HDL-C), fasting blood glucose (FBG), and homocysteine (Hcy) were measured using a fully automated biochemical analyzer (HITACHI-7600-P). All data were collected under standardized conditions following a uniform procedure.

During the period from 2021 to 2022, a whole-cluster random sampling method was employed to randomly select counties in Tibet’s Ngari Prefecture, Naqu, and Shannan cities. Two townships were then randomly selected from each county, from which two villages would be chosen. A total of 520 individuals participated in the questionnaire and physical examination as subjects of this study, and 511 complete questionnaires were recovered. The survey was reviewed and approved by the Medical Ethics Committee of Tibet University, and all study subjects signed an informed consent form.

### Inclusion and Exclusion Criteria

Inclusion criteria: Tibetan residents aged 45 years and over; normal fasting blood glucose level (FBG); no cognitive deficiencies, able to complete the questionnaire independently; voluntary participation in this study.

Exclusion criteria: age<45 years, severe stroke, coronary heart disease, myocardial infarction, people suffering from peripheral vascular, hepatic and renal insufficiency, psychiatric disorders, etc.; people who have recently taken antihypertensive medication.

### Questionnaire

The general information survey was conducted using the questionnaire from the “Prevention and Control of Chronic Diseases in the Tibetan Plateau” program. The questionnaire was divided into three main sections to obtain data about the demographic characteristics, health behaviors, and hypertension-related information. In the first part, demographic characteristics were collected, including the subjects’ age, gender, place of residence (city, rural, or pastoral area), education level (illiterate, primary, middle, or higher degree), employment status (employed, retired, students, unemployed, or herdsmen), and annual income. The second part collected information on health behaviors, including whether subjects currently smoked (yes, one or more per day; no, I quit/never smoked), how much alcohol they consumed in the past month (daily, frequently (3-6 days a week), occasional (1-2 days a week), no, I quit alcohol, or no alcohol), and the frequency of physical exercise (never, 1-2 times a week, 3-5 times a week, or daily), among other questions. The third part collected hypertension-related information, including whether subjects had been diagnosed with hypertension (yes or no) and whether they were currently taking prescription medications for hypertension (yes or no). The survey was administered face-to-face by an investigator and answered by the respondent. The enumerators received training to ensure they had a consistent understanding of the questionnaire.

### Physical Examination

All anthropometric measurements were conducted by trained examiners using standardized methods. Systolic blood pressure (SBP) and diastolic blood pressure (DBP) were measured three times by trained nurses using an OMRON HBP-1300 sphygmomanometer (China), at 5-minute intervals in a seated position after a 5-minute rest. The average of the second and third measurements was used in the analysis. Height and weight were measured with lightweight clothing and without shoes, and the margin of error was controlled to be within ±0.1 cm and ±0.1 kg. Besides, the body mass index (BMI) was calculated by dividing the weight by the square of the height (kg/m^2^).

### Laboratory Assessment

Blood uric acid (UA), total cholesterol (TC), triglycerides (TG), low-density lipoprotein (LDL-C), high-density lipoprotein (HDL-C), fasting blood glucose (FBG), and homocysteine (Hcy) were measured using a fully automated biochemical analyzer (HITACHI-7600-P).

### DNA Extraction and Genotyping

Genomic DNA was extracted from whole blood cells using a QIAamp DNA purification kit (Qiagen, France), and DNA content was quantified using NanoVue Plus®™ (GE Healthcare Life Sciences, France). Genotyping was performed using published TaqMan methods (Applied Biosciences). Genotypes were assigned according to the combinations of the two single-nucleotide polymorphism allelic forms (rs7412 and rs429358) of the APOE gene, known as ε2, ε3 and ε4. Subjects were divided into three groups according to their genotype: ε2 group (ε2/ε3), ε3 group (ε3/ε3, ε2/ε4) and ε4 group (ε3/ε4).

### Covariates Definition

HTN was defined as SBP ≥140 mm Hg and/or DBP ≥90 mm Hg, or current treatment with antihypertensive medications^[15]^. Altitude ranging from 2500 to 4500 m is considered high altitude, and 4500 to 5500 m is considered very high altitude^[16]^. High altitude polycythemia (HAPC) is diagnosed by Hb ≥210 g/L (males) and ≥190 g/L (females)^[17]^. Hyperhomocysteinemia (HHcy) was defined as serum total homocysteine (tHcy) concentration of ≥15 μmol/l^[18]^. High TC was defined as TC ≥6.2 mmol/L and high TG as TG ≥2.3 mmol/L. Low HDL-C was defined as HDL-C <1.0 mmol/L and high LDL-C as LDL-C ≥4.1 mmol/L, those who meet any of the above criteria are considered to dyslipidemia. High Non-HDL-C was defined as Non-HDL-C ≥4.9 mmol/L^[19]^. High FBG was defined as FBG ≥6.1 mmol/L^[20]^. Subjects with BMI 24.0–27.9 kg/m2 was defined as overweight and BMI ≥28.0 kg/m2 as obesity, and overweight or obesity referred to a BMI ≥24.0 kg/m2^[21]^. Criteria for hyperuricemia were SUA levels ≥360 μmol/L in women and 420 μmol/L in men, respectively^[22]^. Current smoking was defined as subjects who reported smoking either every day or some days at the time of survey. Frequent drinking was defined as subjects who reported drinking daily or frequently during the past 30 days preceding the survey.

### Bayesian Network (BN)

The BN model, which usually combines probability theory and graph theory, is one of the probabilistic graphical models that reveal the probabilistic dependence between variables (nodes). The BN model was constructed to further assess the relationship between baseline characteristics and the prevalence of hypertension, as well as the importance of each variable for the prevalence of hypertension. The model was built based on the tree augmented native (TAN) algorithm in the modeling section of SPSS Modeler (version 18.0), and the parameter learning method was chosen as Bayesian adjustment of small cell counts^[23]^. Arrows connecting two nodes indicate that the two random variables are causally or unconditionally independent. If two nodes do not have arrows, the random variables are conditionally independent^[24]^. Based on these advantages of the BN model described above, the interaction of baseline characteristics and lipid profiles of participants with the prevalence of hypertension, and the significance of these aspects for the prevalence of hypertension were further investigated.

### Statistical Analyses

The SPSS statistical software version 26.0 (IBM Inc., USA) was used for data analysis. The measurement data between groups were analyzed by the Chi-square test. Multivariate logistic regression analysis was applied to examine the relationship between Covariates and hypertension. Significant risk factors were incorporated to construct the BN model. Structure learning of BN was conducted using R studio 4.4.1 (R Development Core Team). P < 0.05 was considered statistically significant. The structure learning was realized using the package “bnlearn”. The BN model and BN reasoning were visualized using Netica software (Norsys Sofware Corp., Vancouver, BC, Canada).

## RESULTS

### Baseline Characteristics

This study included 511 participants. 170 hypertensive patients (94 men and 76 women) and 341 controls (139 men and 202 women) were enrolled. We used chi-square tests to explore the differences in each variable between the hypertension and non-hypertension groups. Sex, age, altitude, overweight/obesity, hyperuricaemia, HHcy, HAPC, TG, LDL-C and Non-HDL-C were statistically significant between the two groups (*P* < 0.05), as reflected in Table 1.

**Table 1.**
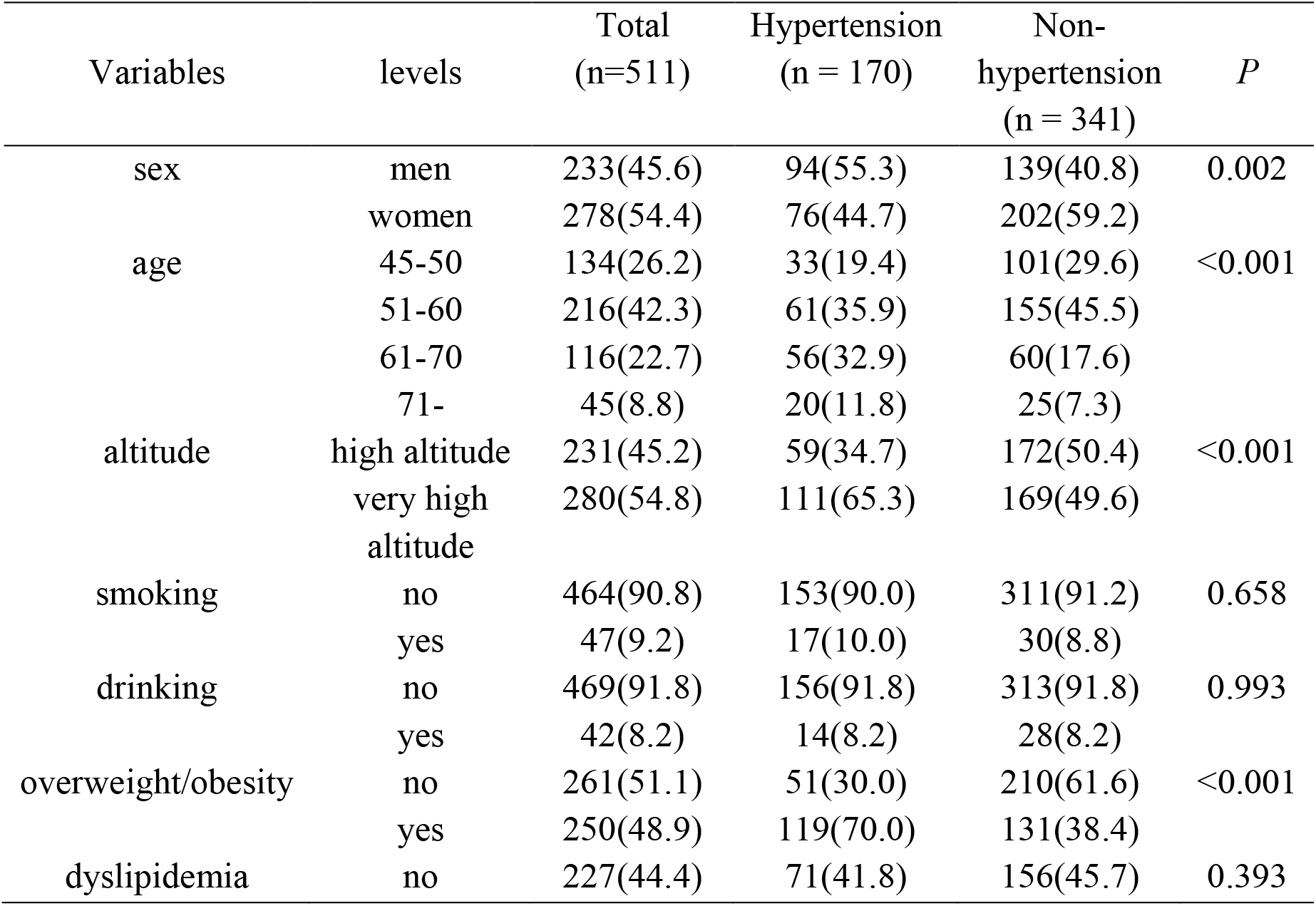
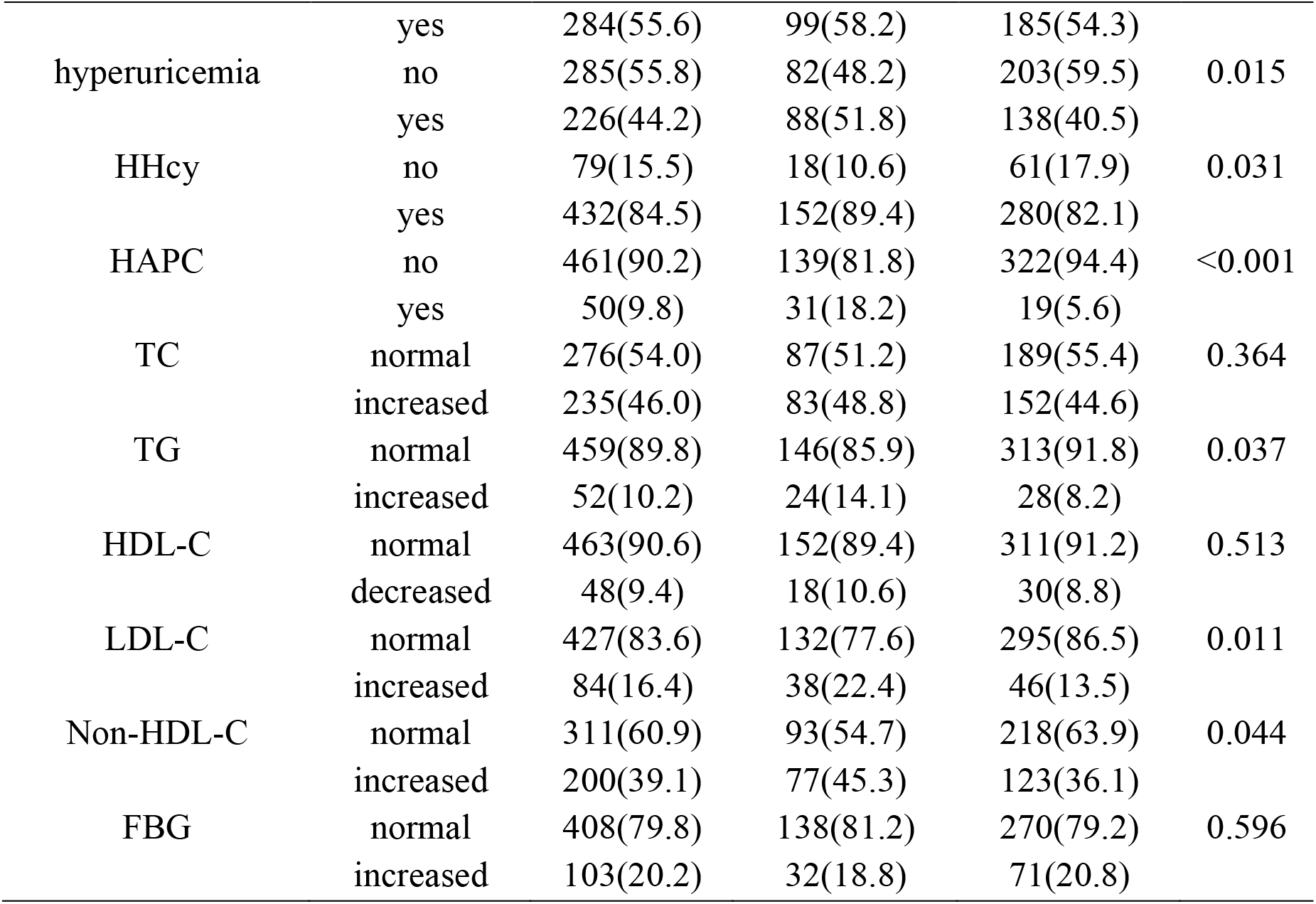
Baseline characteristics of study population.

### Frequencies of APOE and APOE rs7412, rs429358 genotypes

Among all subjects, the frequencies of genotypes ε2/ε3, ε2/ε4, ε3/ε3 and ε3/ε4 were 14.8%,1.8%,75.0%, and 8.4%, respectively. As the results showed, ε3/ε3 was the most common APOE genotype. APOE gene rs7412 locus CC, CT and TT genotype frequencies were 83.4%, 0 and 16.6%, respectively, and rs429358 locus TT, TC and CC genotypes, genotype frequencies were 89.8%, 10.2% and 0. The frequencies of APOE genotypes and rs429358, rs7412 genotypes were compared between hypertensive patients and non-hypertensive controls. The genotype distributions of APOE genotypes and rs429358, rs7412 genotypes in hypertensive patients and controls were consistent with Hardy–Weinberg equilibrium, respectively. It was found that there was no significant difference in the distribution of APOE genotypes and APOE rs7412, rs429358 genotypes between hypertensive patients and controls (*P* > 0.05). (Table 2).

**Table 2.**
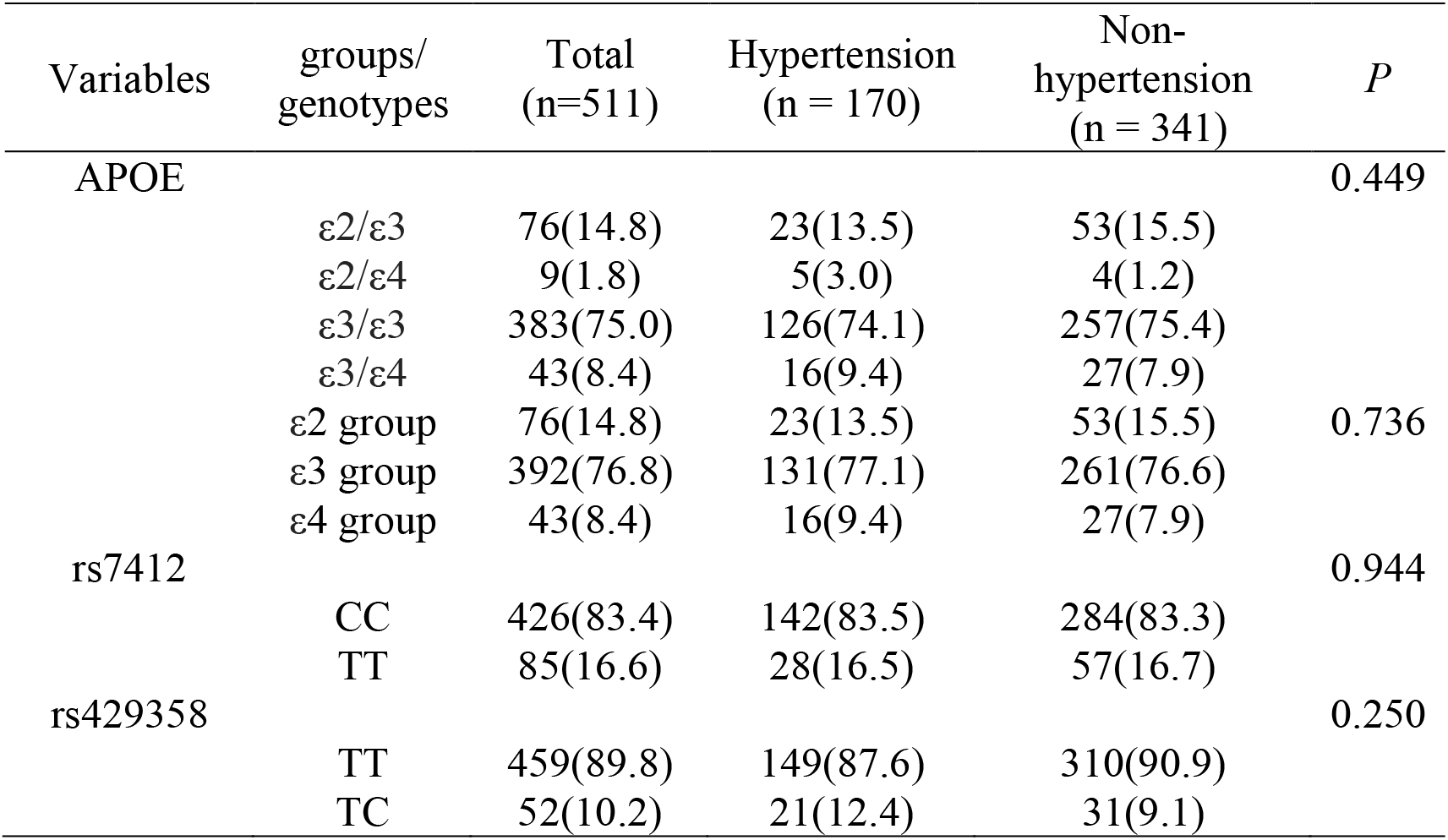
Frequencies of APOE and APOE rs7412, rs429358 genotypes.

### Multivariate Logistic Regression Analysis

We conducted a multivariate logistic regression model with stepwise method (αin = 0.05, αout = 0.10) for risk factors for hypertension, with hypertension presence as the dependent variables, independent variables were those significantly associated with stroke presence in univariate analysis.

The multivariate analysis suggested that hypertension was significantly associated with sex, age, altitude, overweight/obesity, hyperuricaemia, HHcy, HAPC, TG, LDL-Cand Non-HDL-C. (P<0.05, P<0.01). (Table 3)

**Table 3.** Risk factors of hypertension using Logistic regression model.

| Variables | levels | *B* | S.E. | *Wald* | *P* | OR (95% C.I.) |
| --- | --- | --- | --- | --- | --- | --- |
| sex | men |  |  |  |  | Ref |
|  | women | 0.586 | 0.190 | 9.566 | 0.002 | 0.556(0.384, 0.807) |
| age | 45-50 |  |  |  |  | Ref |
|  | 51-60 | 0.186 | 0.251 | 0.549 | 0.459 | 1.204(0.736, 1.970) |
|  | 61-70 | 1.050 | 0.273 | 14.743 | <0.001 | 2.857(1.672, 4.881) |
|  | 71- | 0.895 | 0.361 | 6.159 | 0.013 | 2.448(1.207, 4.966) |
| altitude | high altitude |  |  |  |  | Ref |
|  | very high altitude | 0.650 | 0.194 | 11.196 | <0.001 | 1.915(1.309, 2.801) |
| overweight/obesity | no |  |  |  |  | Ref |
|  | yes | 1.319 | 0.201 | 43.070 | <0.001 | 3.740(2.522, 5.547) |
| hyperuricemia | no |  |  |  |  | Ref |
|  | yes | 0.457 | 0.189 | 5.834 | 0.016 | 1.579(1.090, 2.287) |
| HHcy | no |  |  |  |  | Ref |
|  | yes | 0.610 | 0.287 | 4.526 | 0.033 | 1.840(1.049, 3.226) |
| HAPC | no |  |  |  |  | Ref |
|  | yes | 1.330 | 0.309 | 18.572 | <0.001 | 3.780(2.065, 6.919) |
| TG | normal |  |  |  |  | Ref |
|  | increased | 0.608 | 0.296 | 4.235 | 0.040 | 1.838(1.029, 3.280) |
| LDL-C | normal |  |  |  |  | Ref |
|  | increased | 0.613 | 0.243 | 6.369 | 0.012 | 1.846(1.147, 2.972) |
| Non-HDL-C | normal |  |  |  |  | Ref |
|  | increased | 0.384 | 0.191 | 4.034 | 0.045 | 1.467(1.009, 2.133) |

### Bayesian Network Model

BN was constructed with 11 nodes and 12 directed edges, which is crystal clear in reflecting the risk factors than the Logistic regression model. Directed edges represent probabilistic dependencies between connected nodes. The results suggested sex, age, overweight obesity, HHcy and HAPC represent direct risk factors for hypertension. APOE and APOE rs7412, rs429358 genotypes do not correlate with hypertension. (Figure 1). Based on the maximum likelihood estimation, the common variables predicting hypertension were sex, age, overweight/obesity, HHcy and HAPC (Table 4).

**Table 4.** Conditional probability table.

| age | sex | overweight/obesity | HAPC | HHcy | hypertension |  |
| --- | --- | --- | --- | --- | --- | --- |
|  |  |  |  |  | no | yes |
| 45-50 | man | no | no | no | 0.800 | 0.200 |
| 45-50 | man | yes | no | No | 0.667 | 0.333 |
| 45-50 | man | no | yes | No | 0.500 | 0.500 |
| 45-50 | man | no | no | Yes | 0.783 | 0.217 |
| 45-50 | man | yes | yes | No | 0.500 | 0.500 |
| 45-50 | man | yes | no | yes | 0.556 | 0.444 |
| 45-50 | man | no | yes | yes | 0.500 | 0.500 |
| 45-50 | man | yes | yes | yes | 0.250 | 0.750 |
| 45-50 | woman | no | no | no | 0.938 | 0.625 |
| 45-50 | woman | yes | no | no | 0.692 | 0.308 |
| 45-50 | woman | no | yes | no | 0.500 | 0.500 |
| 45-50 | woman | no | no | yes | 0.867 | 0.133 |
| 45-50 | woman | yes | yes | no | 0.500 | 0.500 |
| 45-50 | woman | yes | no | yes | 0.706 | 0.294 |
| 45-50 | woman | no | yes | yes | 0.500 | 0.500 |
| 45-50 | woman | yes | yes | yes | 0.600 | 0.400 |
| 51-60 | man | no | no | no | 0.667 | 0.333 |
| 51-60 | man | yes | no | no | 0.500 | 0.500 |
| 51-60 | man | no | yes | no | 0.500 | 0.500 |
| 51-60 | man | no | no | yes | 0.821 | 0.179 |
| 51-60 | man | yes | yes | no | 0.500 | 0.500 |
| 51-60 | man | yes | no | yes | 0.614 | 0.386 |
| 51-60 | man | no | yes | yes | 0.800 | 0.200 |
| 51-60 | man | yes | yes | yes | 0.500 | 0.500 |
| 51-60 | woman | no | no | no | 0.643 | 0.357 |
| 51-60 | woman | yes | no | no | 0.429 | 0.571 |
| 51-60 | woman | no | yes | no | 0.667 | 0.333 |
| 51-60 | woman | no | no | yes | 0.863 | 0.137 |
| 51-60 | woman | yes | yes | no | 0.667 | 0.333 |
| 51-60 | woman | yes | no | yes | 0.636 | 0.364 |
| 51-60 | woman | no | yes | yes | 0.667 | 0.333 |
| 51-60 | woman | yes | yes | yes | 0.455 | 0.545 |
| 61-70 | man | no | no | no | 0.800 | 0.200 |
| 61-70 | man | yes | no | no | 0.667 | 0.333 |
| 61-70 | man | no | yes | no | 0.500 | 0.500 |
| 61-70 | man | no | no | yes | 0.438 | 0.562 |
| 61-70 | man | yes | yes | no | 0.500 | 0.500 |
| 61-70 | man | yes | no | yes | 0.286 | 0.714 |
| 61-70 | man | no | yes | yes | 0.333 | 0.667 |
| 61-70 | man | yes | yes | yes | 0.100 | 0.900 |
| 61-70 | woman | no | no | no | 0.727 | 0.273 |
| 61-70 | woman | yes | no | no | 0.500 | 0.500 |
| 61-70 | woman | no | yes | no | 0.500 | 0.500 |
| 61-70 | woman | no | no | yes | 0.840 | 0.160 |
| 61-70 | woman | yes | yes | no | 0.500 | 0.500 |
| 61-70 | woman | yes | no | yes | 0.538 | 0.462 |
| 61-70 | woman | no | yes | yes | 0.500 | 0.500 |
| 61-70 | woman | yes | yes | yes | 0.400 | 0.600 |
| 71- | man | no | no | no | 0.500 | 0.500 |
| 71- | man | yes | no | no | 0.500 | 0.500 |
| 71- | man | no | yes | no | 0.500 | 0.500 |
| 71- | man | no | no | yes | 0.750 | 0.250 |
| 71- | man | yes | yes | no | 0.500 | 0.500 |
| 71- | man | yes | no | yes | 0.375 | 0.625 |
| 71- | man | no | yes | yes | 0.500 | 0.500 |
| 71- | man | yes | yes | yes | 0.333 | 0.667 |
| 71- | woman | no | no | no | 0.667 | 0.333 |
| 71- | woman | yes | no | no | 0.500 | 0.500 |
| 71- | woman | no | yes | no | 0.500 | 0.500 |
| 71- | woman | no | no | yes | 0.556 | 0.444 |
| 71- | woman | yes | yes | no | 0.500 | 0.500 |
| 71- | woman | yes | no | yes | 0.500 | 0.500 |
| 71- | woman | no | yes | yes | 0.250 | 0.750 |
| 71- | woman | yes | yes | yes | 0.333 | 0.667 |

**Figure 1.**
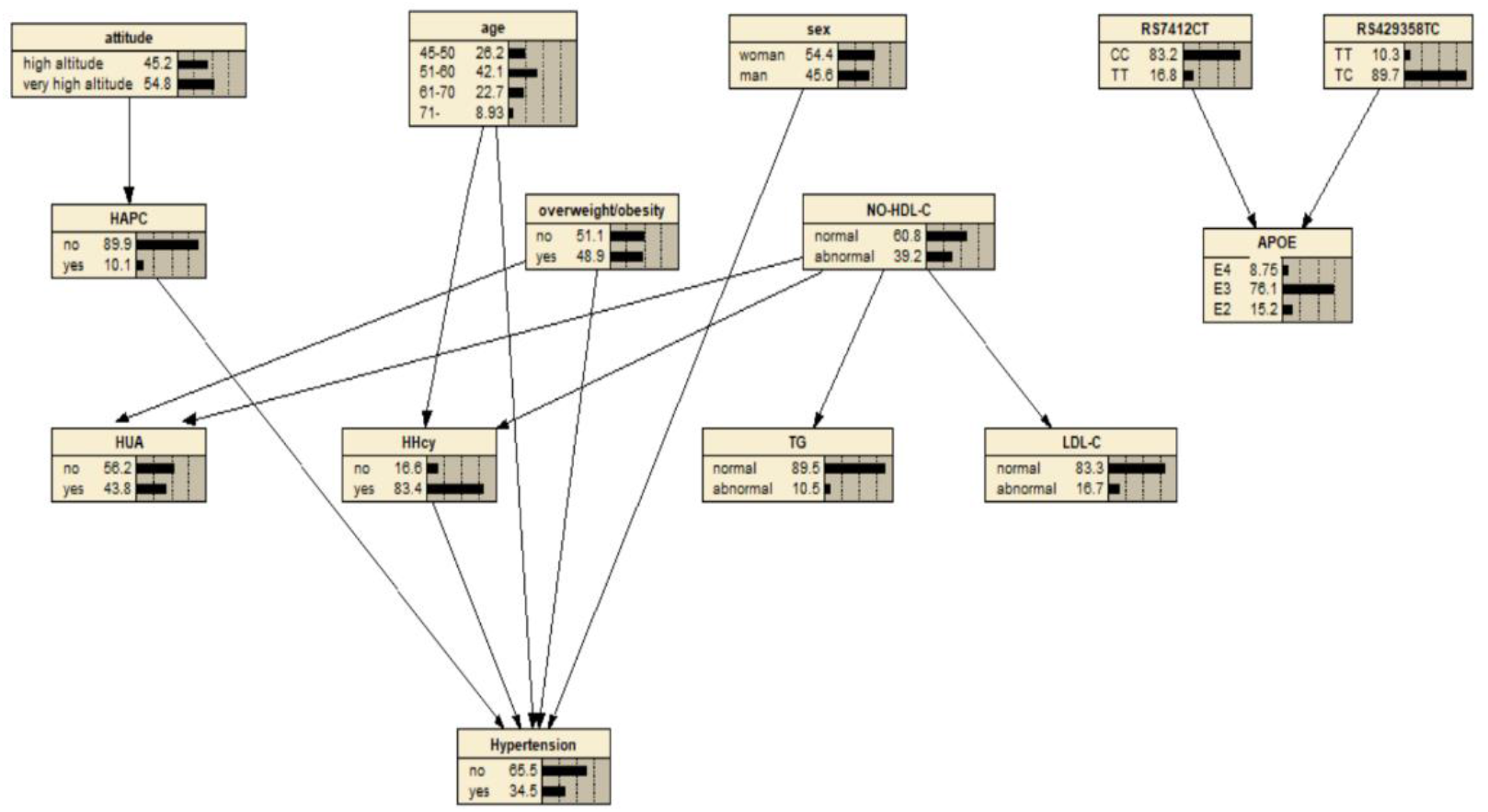
Bayesian network model for hypertension based on TAN algorithm.

### Bayesian Reasoning

Prior probabilities of the variables are presented in Figure 2. The resulting probabilistic model could quantitatively analyse the influence of these factors on hypertension via computing conditional probabilities. If a patient is overweight/obese with HHcy, Bayesian reasoning shows that the risk of hypertension is increased to 45.3%.

**Figure 2.**
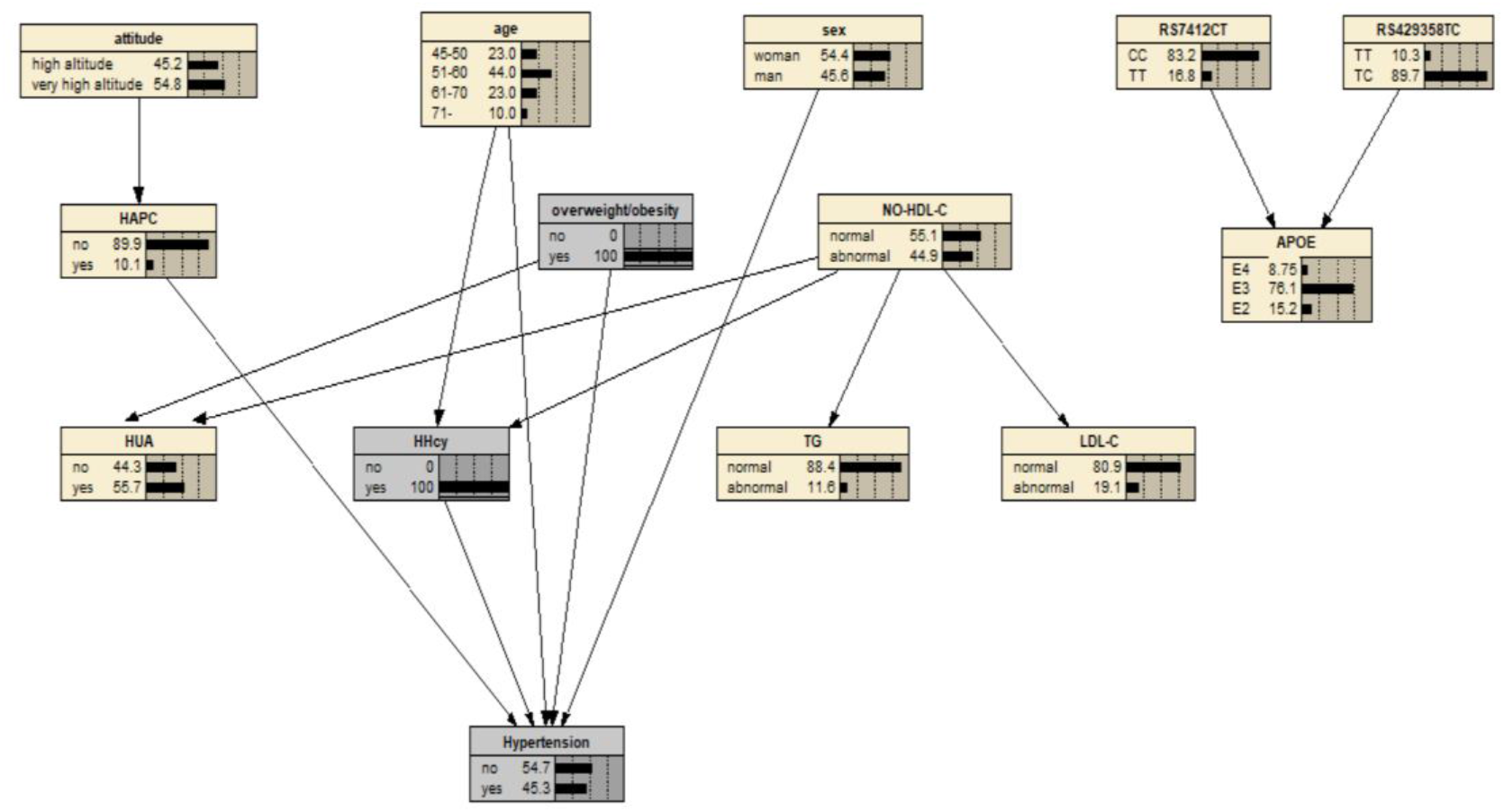
Bayesian networks risk inference for hypertension in middle-aged and elderly Tibetans

## DISCUSSION

### Distribution of APOE and APOE rs7412, rs429358 genotypes in the study population

The results of this study indicated that the frequencies of APOE ε2, ε3 and ε4 alleles were 8.3%, 86.6% and 5.1% in the middle-aged and elderly Tibetan population in Tibet, China. Globally, the ε3 allele is the most common isoform, with frequencies of 85% in Asia, 79% in Europe, 69% in Africa, 82% in North America, and 77% in South America^[25]^. The ε2 and ε4 alleles are less common, accounting for approximately 5%-10% and 10%-15% of human APOE alleles, respectively^[26]^. The ε4 allele is the second most common isoform worldwide, particularly enriched in Central Africa (40%), Oceania (37%), and among Indigenous Australian populations (26%). The distribution of the ε4 allele across Europe and Asia follows a clear north-south gradient, with lower frequencies (<10%) observed in the Mediterranean region and southern China, where the allele frequency in the Chinese population is 6.2%^[27, 28]^. As the most recently evolved subtype, the emergence time of ε2 is difficult to determine, and its global distribution is rare, primarily found in a small portion of African and Oceanic populations^[25]^.

Additionally, the results of this study showed that the proportions of the APOE genotype groups ε2, ε3, and ε4 in the middle-aged and elderly Tibetan population were 14.8%, 76.8%, and 8.4%, respectively. In Beijing^[29]^, the proportions were 12.3%, 70.4%, and 17.3%; in Henan Province^[30]^, they were 15.2%, 73.5%, and 11.3%, in Wuhan^[31]^, they were 13.2%, 70.5%, and 16.3%, and in Chongqing^[32]^, they were 15.3%, 72.2%, and 12.5%. This study found a higher proportion of the common ε3 genotype and a lower proportion of the risky ε4 genotype compared to other regions.

The presence of CC and TT genotypes at the rs7412 C/T locus of the APOE gene in the Tibetan elderly, with frequencies of 83.4% and 16.6%, respectively, and TT and TC genotypes at the rs429358 T/C locus, with frequencies of 89.8% and 10.2%, respectively. Among the Han population in Henan, the rs429358 T/C locus displays TT, TC, and CC genotype frequencies of 87.1%, 12.4%, and 0.5%, respectively. The rs7412 C/T locus shows CC, CT, and TT genotype frequencies of 83.2%, 14.9%, and 1.9%, respectively^[30]^. In the Han population of Guangzhou Province, the rs429358 T/C locus has TT, TC, and CC genotype frequencies of 80.9%, 8.2%, and 0.9%, respectively, and the rs7412 C/T locus has frequencies of 86.4%, 13.0%, and 0.6% for CC, CT, and TT genotypes, respectively^[33]^. This study’s results indicate a mutation frequency of 0% for the CC genotype at the rs429358 T/C locus, and a relatively high mutation frequency of 16.6% for the TT genotype at the rs7412 C/T locus.

### Factors Influencing Hypertension in the middle-aged and elderly Tibetan population

The prevalence of hypertension among middle-aged and elderly Tibetans in China was 33.27%. The logistic regression results indicated that sex, age, altitude, overweight/obesity, hyperuricemia, HHcy, HAPC, TG, LDL-C, and Non-HDL-C were the influencing factors of hypertension. The results suggested sex, age, overweight/obesity, HHcy and HAPC represent direct risk factors for hypertension based on a Bayesian network.

There is a clear association between sex and hypertension^[34]^. Numerous studies have demonstrated that the prevalence of hypertension is higher in men than in women, and these differences may be partly attributed to genetic variations between the sexes, which influence susceptibility and prevalence of hypertension. Additionally, it may be due to sex differences in physiological structures, which lead to significant differences in the regulation of blood pressure homeostasis at the tissue, cellular and molecular levels. Numerous studies have established that age is a significant factor contributing to hypertension^[35]^. As age increases, both the incidence and prevalence of hypertension rise markedly. This study indicates that the risk of hypertension is 2.857 times higher for individuals aged 61-70 and 2.448 times higher for those aged 71 and above, compared to individuals aged 45-50. Although the sample size of the elderly population surveyed decreases with age, the prevalence of hypertension significantly increases. This suggests that the risk of hypertension among Tibetan elderly individuals gradually increases with age.

There is a notable association between elevated blood pressure and increased body mass^[36, 37]^. In overweight/obese populations, factors such as blood viscosity, blood volume, peripheral vascular resistance, and cardiac output increase, all of which are related to hypertension. Additionally, Homocysteine (Hcy) levels are also closely linked to hypertension, for every 10 μmol/L increase in Hcy, systolic and diastolic blood pressure increase by 1 mmHg and 1.4 mmHg, respectively^[38]^. Elevated serum Hcy levels stimulate smooth muscle cell proliferation and induce endothelial cell damage, which compromises vascular integrity and contributes to increased blood pressure^[39]^.

High Altitude Pulmonary Edema (HAPC), characterized by excessive compensatory erythrocyte proliferation due to hypoxia, is a common chronic altitude sickness that typically occurs in regions above 3,000 meters. It can lead to increased blood viscosity, microcirculatory disorders, thrombus formation, widespread organ damage, and sleep disturbances^[40]^. The incidence of HAPC is positively correlated with altitude, as altitude increases, hypoxia becomes more severe, resulting in a higher prevalence of chronic altitude sickness. In HAPC patients, elevated blood pressure is primarily characterized by increased diastolic pressure, likely related to increased blood viscosity and blood volume^[41]^. Consistent with the Bayesian network model, altitude indirectly affects hypertension.

The results of this study indicated no correlation between APOE genotypes and APOE rs7412, rs429358 genotypes, and the prevalence of hypertension in middle-aged and elderly Tibetan population in Tibet. This finding is consistent with studies conducted by several researchers both domestically and internationally^[42-45]^, and may be related to the sample size of this study. Conversely, other research suggests that APOE gene is closely related to blood pressure, the potential mechanism may be that APOE gene affects blood pressure by regulating lipid metabolism^[46]^. Notably, the ApoE ε4 allele has previously been identified as a risk factor for hypertension^[47]^.

### Strengths and Limitations

In this study, we investigated APOE genotypes and APOE rs7412, rs429358 genotypes in a middle-aged and elderly Tibetan population in Tibet to explore the frequency distribution and the relationship with hypertension. However, there are several limitations, due to the sparsely populated and difficult conditions in Tibet, resulting in small sample sizes in some high-altitude and very high-altitude areas, and the cross-sectional study was unable to determine a causal relationship. Further expansion of the sample size is needed to improve the application value of the model.

## Conclusions

The prevalence of hypertension among middle-aged and elderly Tibetans in China was 33.27%. The percentage of APOE ε2, ε3 and ε4 alleles were 8.3%, 86.6% and 5.1%. The proportion of APOE genotype ε2, ε3 and ε4 groups were 14.8%, 76.8% and 8.4%, respectively. In the Tibetan elderly population, the frequencies of the CC and TT genotypes at the rs7412 C/T locus of the APOE gene were 83.4% and 16.6%, respectively. For the rs429358 T/C locus, the frequencies of the TT and TC genotypes were 89.8% and 10.2%. The logistic regression analysis revealed that sex, age, altitude, overweight/obesity, hyperuricemia, HHcy, HAPC, TG, LDL-C, and Non-HDL-C were the influencing factors of hypertension. Sex, age, overweight obesity, HHcy and HAPC represent direct risk factors for hypertension based on a Bayesian network.

### Perspectives

In this study, APOE genotype and APOE rs7412, rs429358 genotypes were not genetically correlated with hypertension in middle-aged and elderly Tibetan populations in Tibet. This may be related to the small sample. Further expansion of the sample size is needed to improve the application value of the study. Sex, age, overweight obesity, HHcy and HAPC represent direct risk factors for hypertension based on a Bayesian network, which provided a scientific basis for the early prevention of hypertension among middle-aged and elderly Tibetan populations in Tibet.

## Data Availability

The datasets that support the findings of this study are available from Medical College of Tibet University but are restricted to the availability of these data, which were used under license for the current study, and so are not publicly available. Data are however available from the authors upon reasonable request and with permission of Medical College of Tibet University. If someone wants to request data from this study, please contact the corresponding author.

## Non-standard Abbreviations and Acronyms

HTN: hypertension
SBP: systolic blood pressure
DBP: diastolic blood pressure
UA: Blood uric acid
TC: total cholesterol
TG: triglycerides
LDL-C: low-density lipoprotein cholesterol
HDL-C: high-density lipoprotein cholesterol
FBG: fasting blood glucose
Hcy: homocysteine
HHcy: hyperhomocysteinemia
BMI: Body Mass Index
HAPC: High Altitude Pulmonary Edema
OR: Odds ratio
CI: Confidence interval

## Acknowledgements

We would like to express our gratitude to all those who helped us during the writing of this research. We extend our deepest gratitude to Professor Hai Xiong, who provided us with valuable suggestions.

## Affiliations

Medical College of Tibet University, Lhasa, Tibet Autonomous Region, China (Y.Z., J.Z., D.Q., H.C., H.X.,). Department of Cardiology, West China School of Public Health and West China Fourth Hospital Sichuan University (Y.Y.,) and Sichuan University (J.Y.,), Chengdu, Sichuan Province, China.

## Ethics approval and consent to participate

The Medical ethics committee of Tibet University has approved the study, and all participants provided written informed consent.

